# ChatGPT for automating lung cancer staging: feasibility study on open radiology report dataset

**DOI:** 10.1101/2023.12.11.23299107

**Authors:** Yuta Nakamura, Tomohiro Kikuchi, Yosuke Yamagishi, Shouhei Hanaoka, Takahiro Nakao, Soichiro Miki, Takeharu Yoshikawa, Osamu Abe

## Abstract

**Objectives:** CT imaging is essential in the initial staging of lung cancer. However, free-text radiology reports do not always directly mention clinical TNM stages. We explored the capability of OpenAI’s ChatGPT to automate lung cancer staging from CT radiology reports.

**Methods:** We used MedTxt-RR-JA, a public de-identified dataset of 135 CT radiology reports for lung cancer. Two board-certified radiologists assigned clinical TNM stage for each radiology report by consensus. We used a part of the dataset to empirically determine the optimal prompt to guide ChatGPT. Using the remaining part of the dataset, we (i) compared the performance of two ChatGPT models (GPT-3.5 Turbo and GPT-4), (ii) compared the performance when the TNM classification rule was or was not presented in the prompt, and (iii) performed subgroup analysis regarding the T category.

**Results:** The best accuracy scores were achieved by GPT-4 when it was presented with the TNM classification rule (52.2%, 78.9%, and 86.7% for the T, N, and M categories). Most ChatGPT’s errors stemmed from challenges with numerical reasoning and insufficiency in anatomical or lexical knowledge.

**Conclusions:** ChatGPT has the potential to become a valuable tool for automating lung cancer staging. It can be a good practice to use GPT-4 and incorporate the TNM classification rule into the prompt. Future improvement of ChatGPT would involve supporting numerical reasoning and complementing knowledge.

**Clinical relevance statement:** ChatGPT’s performance for automating cancer staging still has room for enhancement, but further improvement would be helpful for individual patient care and secondary information usage for research purposes.

**Key points:**

- ChatGPT, especially GPT-4, has the potential to automatically assign clinical TNM stage of lung cancer based on CT radiology reports.
- It was beneficial to present the TNM classification rule to ChatGPT to improve the performance.
- ChatGPT would further benefit from supporting numerical reasoning or providing anatomical knowledge.

**Graphical abstract:** 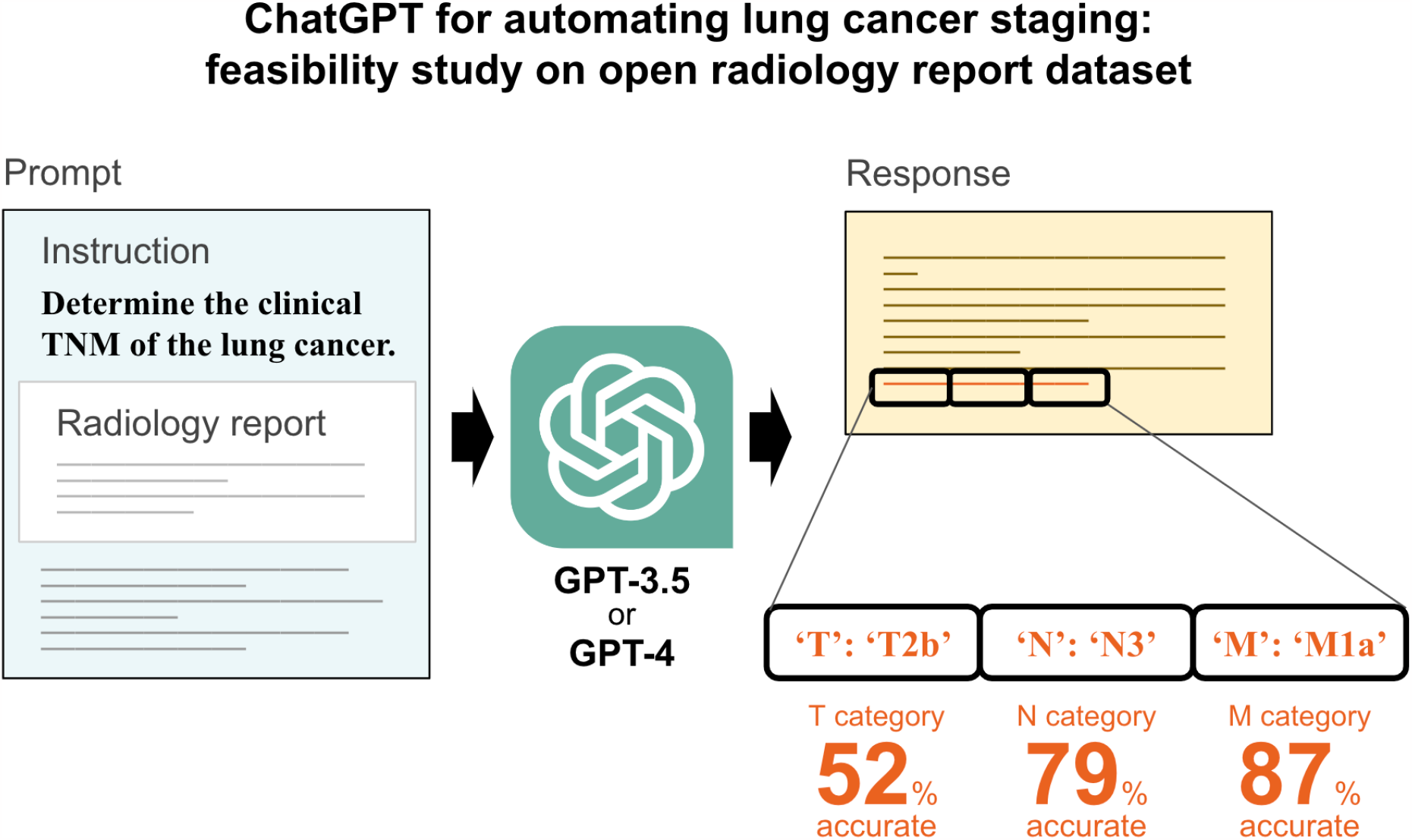

## Introduction

Lung cancer is a primary cause of cancer-related mortalities around the world [1]. The TNM classification for lung cancer [2] is an essential tool for individual patient care and clinical research. However, the current TNM classification rule is complicated, making it difficult even for radiologists to memorize it all. As a result, although tumor characteristics are described in unstructured radiology reports, clinical TNM stage (cTNM) is not always explicitly mentioned [3]. Physicians often have to scrutinize radiology reports to extract relevant staging information for clinical or research purposes.

To alleviate workloads, several natural language processing (NLP) techniques have been proposed to automate staging from unstructured radiology reports. While their potential has been demonstrated in existing studies, they often depended on meticulously organized rules and vocabularies [4–6] or large training datasets [7–9].

OpenAI’s ChatGPT [10] has opened a new era where people can use NLP techniques through a conversational interface. Performance for various clinical tasks has been reported, including medical license examinations [11–16], board certification examinations [17–21], and image interpretation quizzes [22, 23]. Notably, ChatGPT can solve various tasks by simply providing it with an instruction (or “prompt”) written in a natural language without the need for further task-specific preparations. Given that cancer staging is a complex task demanding extensive clinical knowledge, assessing ChatGPT’s capability in staging is worthwhile. To the best of our knowledge, no previous researches have applied ChatGPT to automated cancer staging from unstructured radiology reports.

The purpose of this study is to evaluate ChatGPT’s potential for automating lung cancer staging and explore optimal prompts to guide ChatGPT effectively.

## Materials and Methods

### Ethical considerations

We used an existing public dataset containing no personal health information [24]. Hence, approval by the institutional review board was waived for this retrospective feasibility study.

### Dataset

#### Radiology reports

In this study, we utilized MedTxt-RR-JA [24], an open dataset consisting of 135 CT radiology reports for lung cancer. This dataset is based on 15 cases of lung cancer at varying stages on a public educational website (Radiopaedia^1^), with each case independently diagnosed by nine radiologists. Thus, the dataset is free from privacy concerns. All the radiology reports are written in Japanese using a free narrative style, without structured sections such as “impression” or “conclusion”.

82 of the 135 (60.7%) radiology reports directly mentioned cTNM, which could lead to overestimating ChatGPT’s performance. We masked all such explicit mentions by manually substituting them with the placeholder “##MASK##.” We made no other modifications to the radiology reports.

#### Gold standard

Two board-certified radiologists assigned the gold standard for cTNM to each radiology report. The radiologists had six and seven years of experience, respectively, in diagnostic radiology. They independently reviewed the masked radiology reports and assigned cTNM. They did not refer to the direct mentions of cTNM in the original (unmasked) radiology reports, nor to the original CT images on Radiopaedia. After completing the independent assignment, the inter-observer agreement was measured using Cohen’s kappa coefficient. Any discrepancy was resolved through subsequent discussions.

The gold standard was based on the 8th edition of the TNM classification rule by the Japan Lung Cancer Society [25], which closely aligns with the 8th edition of the AJCC/UICC TNM classification rule [2, 26]. For the T category, the options were T0, Tis, T1mi, T1a, T1b, T1c, T2a, T2b, T3 and T4; for the N category, N0, N1, N2, and N3; and for the M category, M0, M1a, M1b, and M1c.

#### Data split

The dataset was randomly split into two subsets: the *development set* containing 45 radiology reports from five cases, and the *test set* containing 90 radiology reports from ten cases. This case-based split ensured that radiology reports for the same case did not appear in both sets.

### Experiment 1: GPT-3.5 Turbo vs GPT-4

#### Overview

We compared the performance of two ChatGPT models, GPT-3.5 Turbo [27] and GPT-4 [10]. We used the model snapshots on June 13, 2023. Our experiments were conducted from September 28 to October 11, 2023.

First, using the development set, we searched for the optimal prompt to instruct ChatGPT. After finalizing the prompt, we used the test set to examine the performance of GPT-3.5 Turbo and GPT-4.

We communicated with ChatGPT through OpenAI’s application programming interface (API). To repeatedly evaluate ChatGPT’s performance, we wrote a custom Python (version 3.9.10) script using the OpenAI library version 0.28.1. The *temperature* parameter passed to the API was set to zero, ensuring that ChatGPT would always produce the same responses to identical prompts [28]. Given this deterministic feature, we conducted just one trial per radiology report.

#### Prompt Optimization

First, we experimentally determined the optimal prompt to be used in subsequent analysis. We repeatedly tried various prompts containing each radiology report in the development set to explore the optimal prompt that could maximize the staging performance. After empirically trying more than 100 prompts, we found that simplistic prompts sometimes led ChatGPT to refuse to answer, and ended up with providing a comprehensive prompt with detailed instructions.

As shown in Figure 2, the finalized prompt had multiple components: (i) a primary instruction, (ii) the radiology report to be analyzed, (iii) stepwise instructions described below, and (iv) the TNM classification rule.

**Figure 1:**
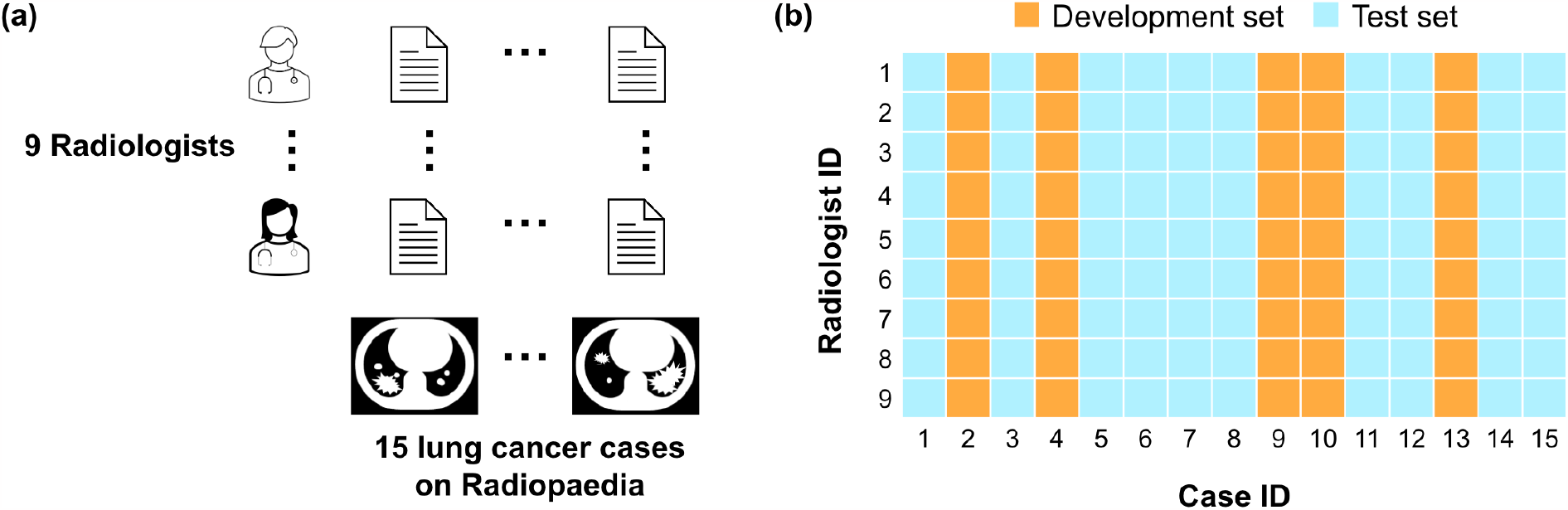
Dataset in this study. (a) We used an existing open dataset [24], contributed by nine radiologists who independently diagnosed fifteen cases on Radiopaedia. (b) We split the dataset into the development and test sets on a case basis.

**Figure 2:**
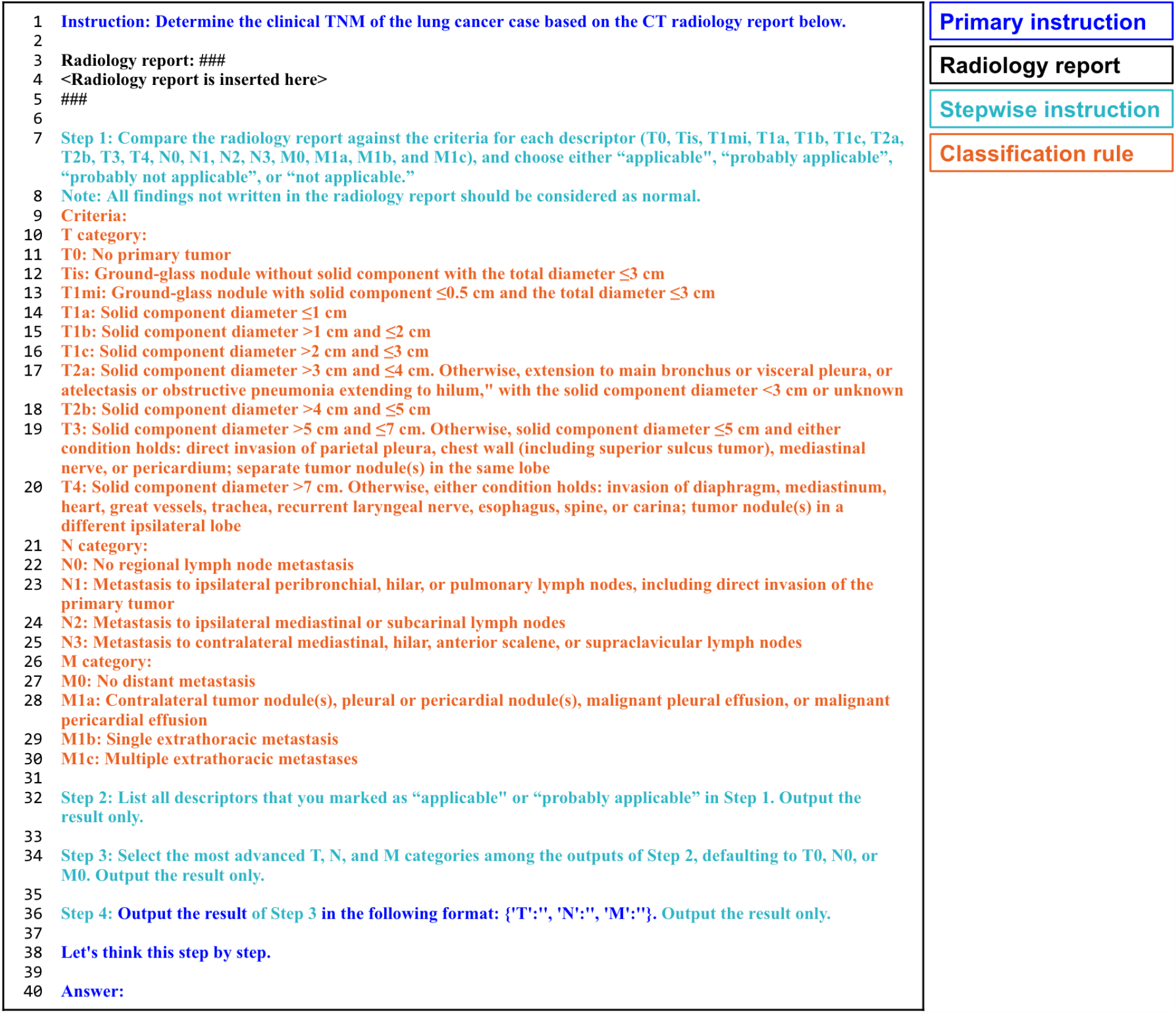
The prompt sent to ChatGPT to automate lung cancer staging (English translation from the original Japanese edition). ChatGPT will respond, continuing from “Answer:” writing its intermediate inference and answer. The line numbers to the left of the prompt text are only for display here.

The stepwise instructions guided ChatGPT to solve the problem in three steps. First, we told ChatGPT to determine whether the radiology report agreed with the criterion for each option (i.e., T0, Tis, T1mi, etc.). Next, we instructed ChatGPT to list all the matches and pick the most progressed one for the T, N, and M categories, respectively. Finally, we asked ChatGPT to output the final staging result in a JSON-like format such as `{‘T’: ‘T2a’, ‘N’: ‘N1’, ‘M’: ‘M1b’}`.

We included in the prompt a tailored short version of the TNM classification rule [25]. We also told ChatGPT to default to T0, N0, or M0 without leaving blanks when it failed to determine the answer. We encouraged ChatGPT to explain the reasoning process [29]. The prompt was written entirely in Japanese.

Our experiment was based on a *zero-shot* setting. That is, ChatGPT relied on its inherent clinical knowledge and instructions in the prompt but used no prior training data or examples.

#### Evaluation

Following previous researches [4–6], we calculated the accuracy score, or the proportion of correct answers, for the T, N, and M categories.

We extracted staging results from the ChatGPT response using regular expressions and only accepted answers following the JSON-like format as valid. We allowed the omission of capitals, such as ‘2a’ instead of ‘T2a’, but regarded any other variation as incorrect. Any discrepancies in subdivisions, such as T2a versus T2b, were also considered incorrect.

#### Statistical test

We performed the McNemar test with a continuity correction to compare the performance of GPT-3.5 Turbo and GPT-4 on the test set. The McNemar test was based on contingency tables to count correct and incorrect answers by each model. The significance standard was set to *p* = 0.05. All the statistical analyses were performed using R (version 4.1.3; R Foundation for Statistical Computing, Vienna, Austria).

### Experiment 2: Ablation study

We added an experiment to determine whether it was beneficial to provide ChatGPT with the TNM classification rule in the prompt. We conducted the same study as Experiment 1 but with the TNM classification rule omitted from the prompt. We conducted the McNemar test to examine the performance change from Experiment 1.

### Experiment 3: Subgroup analysis for the T category

The classification rule for the T category is more complex than those for the N and M categories, as it involves three independent factors: (i) tumor size, (ii) tumor extension, and (iii) presence of satellite lesions. We conducted an additional subgroup analysis focusing on the T category to identify the factor that most affected ChatGPT’s performance.

For each radiology report in the test set, we identified which of the three factors was decisive in determining the T category. For instance, in a T3 case where an 18 mm primary tumor invades the chest wall without any satellite lesions, the tumor extension was considered the T-determining factor.

Based on the prediction results in Experiment 1, we computed accuracy scores for each of the three T-determining factors. Subsequently, we performed binomial tests to ascertain if the differences in accuracy scores among the three T-determining factors were statistically significant. We used a corrected significance standard of *p* = 0.05*/*3 = 0.0166 … to account for multiple comparisons according to the Bonferroni correction.

## Results

### Dataset

Table 1 shows the distribution of the T, N, and M categories in our gold standard. No radiology reports were labeled as T0, T1a, or T1b. Cohen’s kappa coefficients of inter-observer agreements of the T, N, and M categories were 0.97, 0.98, and 0.95, suggesting near-perfect agreement between the two radiologists.

**Table 1:** The population of the development and test sets.

|  | Total |  | Development set |  | Test set |  |
| --- | --- | --- | --- | --- | --- | --- |
| Tis | 6 | (4.4%) | 4 | (8.9%) | 2 | (2.2%) |
| T1mi | 12 | (8.9%) | 5 | (11.1%) | 7 | (7.8%) |
| T1c | 9 | (6.7%) | 0 | (0.0%) | 9 | (10.0%) |
| T2a | 23 | (17.0%) | 9 | (20.0%) | 14 | (15.6%) |
| T2b | 10 | (7.4%) | 1 | (2.2%) | 9 | (10.0%) |
| T3 | 24 | (17.8%) | 7 | (15.6%) | 17 | (18.9%) |
| T4 | 51 | (37.8%) | 19 | (42.2%) | 32 | (35.6%) |
| Total | 135 | (100.0%) | 45 | (100.0%) | 90 | (100.0%) |
| N0 | 68 | (50.4%) | 24 | (53.3%) | 44 | (48.9%) |
| N1 | 8 | (5.9%) | 2 | (4.4%) | 6 | (6.7%) |
| N2 | 28 | (20.7%) | 11 | (24.4%) | 17 | (18.9%) |
| N3 | 31 | (23.0%) | 8 | (17.8%) | 23 | (25.6%) |
| Total | 135 | (100.0%) | 45 | (100.0%) | 90 | (100.0%) |
| M0 | 97 | (71.9%) | 35 | (77.8%) | 62 | (68.9%) |
| M1a | 8 | (5.9%) | 1 | (2.2%) | 7 | (7.8%) |
| M1b | 4 | (3.0%) | 0 | (0.0%) | 4 | (4.4%) |
| M1c | 26 | (19.3%) | 9 | (20.0%) | 17 | (18.9%) |
| Total | 135 | (100.0%) | 45 | (100.0%) | 90 | (100.0%) |

### Experiment 1: GPT-3.5 Turbo vs GPT-4

Table 2 summarizes the performance of GPT-3.5 Turbo and GPT-4 for automating lung cancer staging. On the test set, GPT-3.5 Turbo marked the accuracy scores of 37.8%, 68.9%, and 67.8% for the T, N, and M categories, respectively. With the same prompt, GPT-4 surpassed GPT-3.5 Turbo with an accuracy score of 52.2% (*p* = 0.03), 78.9% (*p* = 0.08), and 86.7% (*p <* 0.01) for the T, N, and M categories. GPT-4 outperformed GPT-3.5 Turbo with statistical significance for the T and M categories.

**Table 2:**
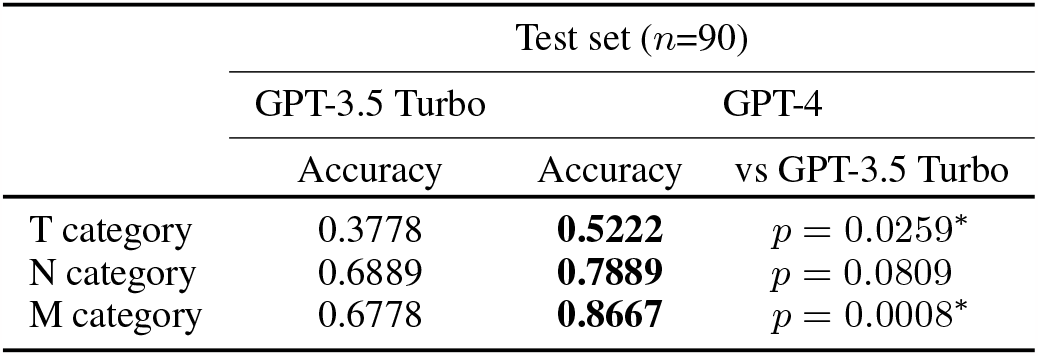
The accuracy scores, with the best values shown in **bold**. ^*^Statistical significance in McNemar test.

| | Test set ($n=90$) | | |
| --- | --- | --- | --- |
|  | GPT-3.5 Turbo | GPT-4 |  |
|  | Accuracy | Accuracy | vs GPT-3.5 Turbo |
| T category | 0.3778 | **0.5222** | $p = 0.0259^*$ |
| N category | 0.6889 | **0.7889** | $p = 0.0809$ |
| M category | 0.6778 | **0.8667** | $p = 0.0008^*$ |

### Experiment 2: Ablation study

Table 3 shows the ablation study result. The omission of the TNM classification rule from the prompt negatively affected both GPT-3.5 Turbo and GPT-4. The decrease in the accuracy score was statistically significant for the T and N categories.

**Table 3:** The result of ablation study, with the best values shown in bold. The reference score is the same as those appears in Table 2. ^*^Statistical significance in McNemar test.

| | Test set ($n=90$) | | |
| --- | --- | --- | --- |
|  | GPT-3.5 Turbo |  |  |
|  | Classification rule (+) | (-) |  |
|  | Accuracy (Reference) | Accuracy | vs Reference |
| T category | **0.3778** | 0.2111 | $p = 0.0119^*$ |
| N category | **0.6889** | 0.5111 | $p = 0.0101^*$ |
| M category | 0.6778 | **0.7556** | $p = 0.7728$ |
|  | GPT-4 |  |  |
|  | Classification rule (+) | (-) |  |
|  | Accuracy (Reference) | Accuracy | vs Reference |
| T category | **0.5222** | 0.3778 | $p = 0.0088^*$ |
| N category | **0.7889** | 0.6333 | $p = 0.0012^*$ |
| M category | **0.8667** | 0.8556 | $p = 1.0000$ |

### Experiment 3: Subgroup analysis for the T category

Tumor size, tumor extension, and satellite lesion were the T-determining factors in the 43, 43, and 15 radiology reports in the test set.

Figure 3 shows the result of the subgroup analysis for the T category. For the radiology reports in which the tumor size, the tumor extension, and the presence of satellite lesions were the T-determining factors, the corresponding accuracy scores were 21%, 56%, and 67%, respectively, for GPT-3.5 Turbo, and 51%, 60%, and 80% for GPT-4.

**Figure 3:**
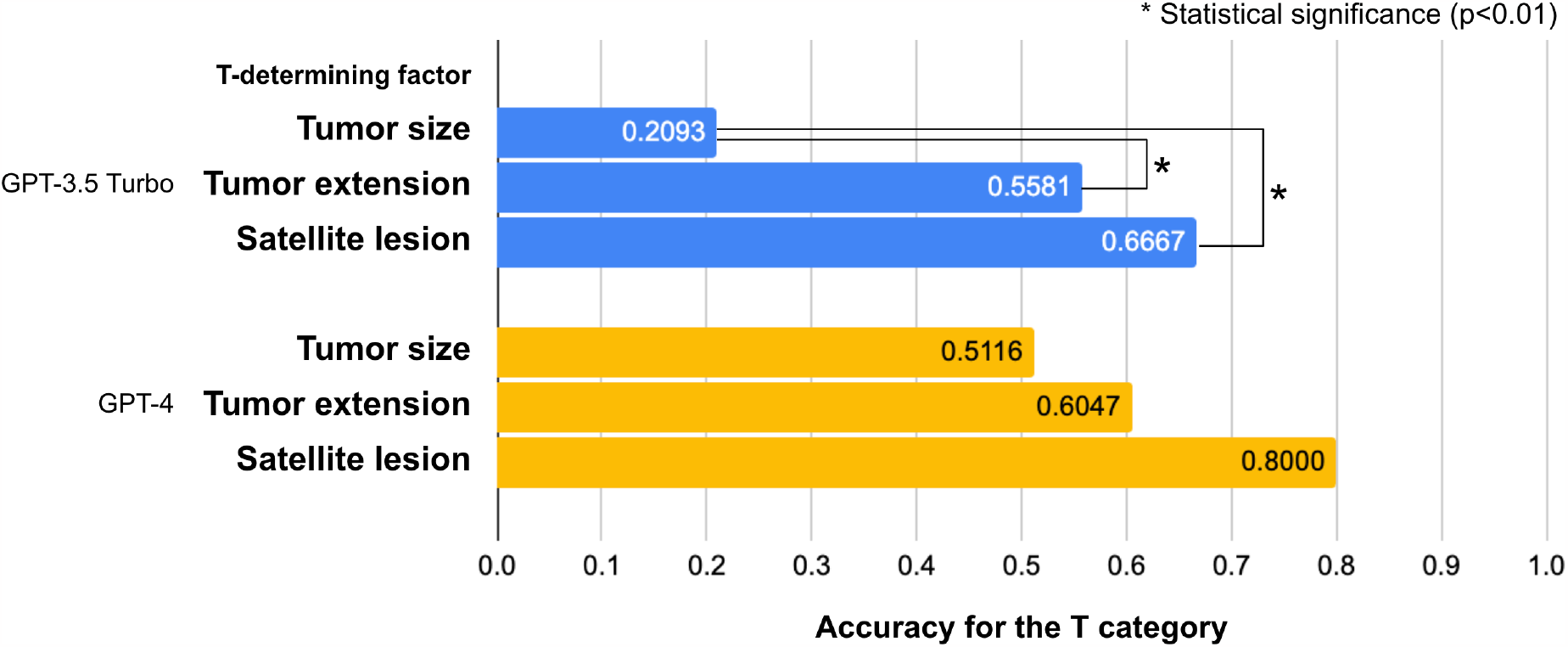
Results of the subgroup analysis for the T category. On the test set, accuracy scores set were calculated separately for different T-determining factors.

The accuracy score was lower in the radiology reports in which the tumor size was the T-determining factor than for the tumor extension and satellite lesion. In particular, when GPT-3.5 Turbo was used, the differences were statistically significant.

## Discussion

We demonstrated that GPT-4 accurately determined the T, N, and M categories of 52%, 79%, and 87% of lung cancer CT radiology reports written in Japanese. To the best of our knowledge, this is the first study that investigated the possibility of ChatGPT for automating cancer staging. Although imperfect, it is noteworthy that this performance was achieved with no prior training specific to this study.

Our experiments showed that GPT-4 was superior to GPT-3.5 Turbo for automating lung cancer staging. The GPT-4’s superiority is consistent with previous researches on medical license examinations [13, 15], as well as on other tasks requiring specialized knowledge and complex reasoning [10].

Our ablation study revealed that ChatGPT needed the TNM classification rule incorporated into the prompt. Although ChatGPT has been reported to pass some board certification examinations [19, 21], its inherent knowledge of the TNM classification rule may not be complete.

Our subgroup analysis implies that ChatGPT’s performance for the T category may be hindered by its numerical comparison ability; it often gave an incorrect T category when the tumor size was the determining factor, even though the criteria were explicitly included in the prompt.

Table 4 summarizes previously suggested methods for automating lung cancer staging from radiology reports. Rule-based methods achieved an accuracy score of around 0.9 for the T and N categories [4–6], and a deep learning method marked the accuracy score of 0.93 for the M category [7]. ChatGPT appears to lag behind existing methods for the T and N categories but seems more competent for the M category. However, direct comparison with previous researches is challenging due to the difference in dataset characteristics; unlike previous researches, the MedTxt-RR-JA dataset [24] does not reflect the real-world population, as Radiopaedia includes various lung cancer cases encompassing a wide range of imaging findings.

**Table 4:**
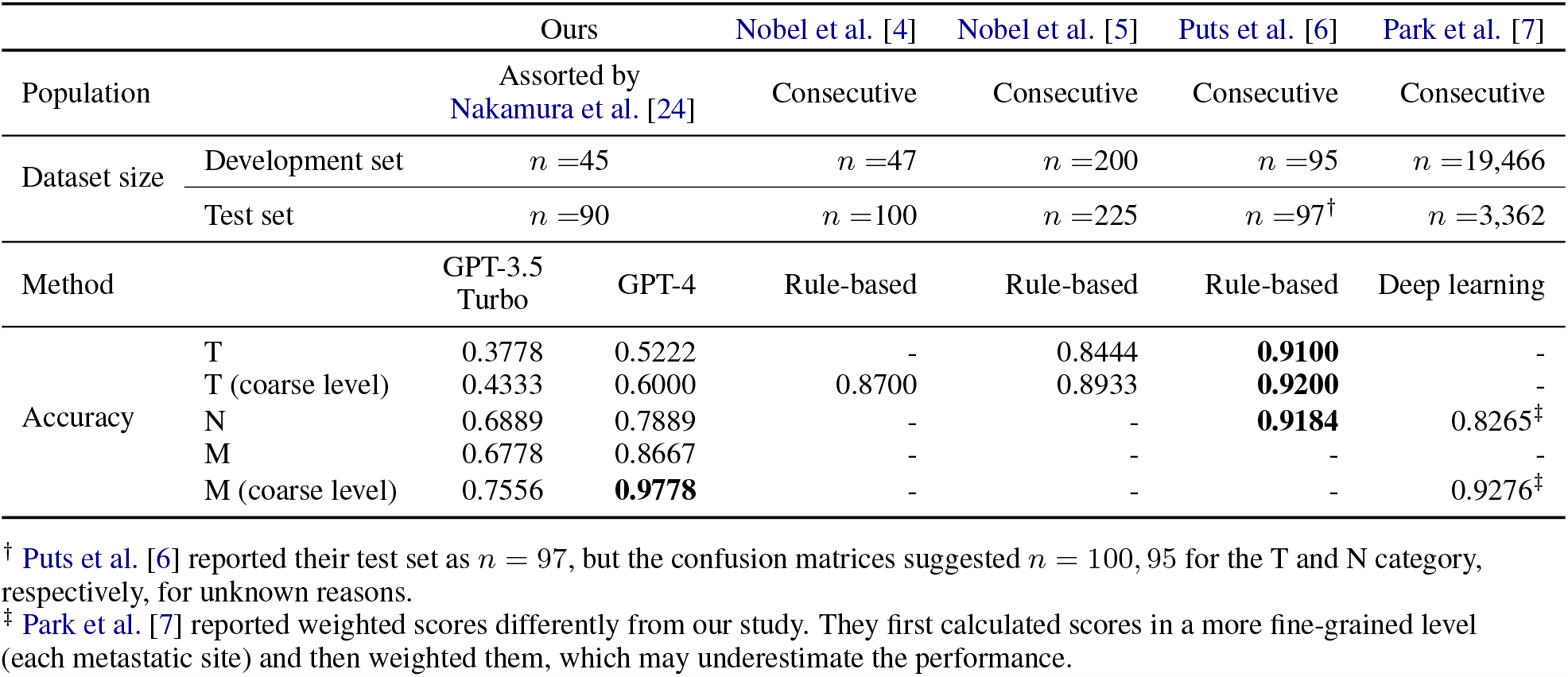
Comparison with previous researches, with the best values shown in bold. The coarse level accuracy scores were calculated by allowing discrepancies among T1mi, T1a, T1b, and T1c; T2a and T2b; and M1a, M1b, and M1c.

Our study had several limitations. First, due to budgetary and time constraints, we optimized the prompt by repeatedly evaluating the accuracy score using only GPT-3.5 Turbo; the finalized prompt might not have been optimal for GPT-4. Second, more aggressive “prompt-engineering” might have yielded an even better performance. We decided not to optimize the prompt excessively to be overly detailed or disease-specific; instead, we only included the TNM classification rule in the prompt. Including disease-specific diagnostic tips in the prompt might have been effective, but we chose not to do so as it might compromise the generalizability of our suggested method. Lastly, we conducted experiments in a *zero-shot* setting. Including several examples of a question and its expected response in the prompt is a well-known strategy for obtaining better responses (known as *few-shot*), and this will be a subject of future research.

In conclusion, ChatGPT may serve as a valuable tool to determine cTNM from radiology reports solely through the use of an optimized prompt, without any specific training. It was effective to use GPT-4 and include the TNM classification rule in the prompt. Future improvement of ChatGPT would involve supporting numerical reasoning and complementing knowledge.

## Data Availability

All data produced are available online at https://sociocom.naist.jp/medtxt/rr/

## Abbreviations and acronyms

cTNM: clinical TNM stage
NLP: natural language processing
API: application programming interface

## Acknowledgements

The Department of Computational Diagnostic Radiology and Preventive Medicine is sponsored by HIMEDIC, Inc. and Siemens Japan KK.

In preparing this manuscript, the authors used ChatGPT-4 to refine the English prose. Following this, the authors carefully reviewed and revised the material as necessary, ensuring full responsibility for the published content.

## Footnotes

1 https://radiopaedia.org/

